# Methylenetetrahydrofolate reductase polymorphic variants in rectal cancer: significance for cancer risk and response to chemoradiotherapy

**DOI:** 10.1101/2023.09.21.23295916

**Authors:** Aleksandra Stanojevic, Jelena Spasic, Mladen Marinkovic, Suzana Stojanovic-Rundic, Radmila Jankovic, Ana Djuric, Jerome Zoidakis, Remond J.A. Fijneman, Sergi Castellvi-Bel, Milena Cavic

## Abstract

**Background:** Methylenetetrahydrofolate reductase (MTHFR) small nucleotide polymorphisms (SNPs) have been suggested as risk, prognostic, and predictive factors for colorectal cancer in various populations, but have not been validated so far. The aim of this study was to analyze the association of *MTHFR* C677T (*rs1801133*) and A1298C (*rs1801131*) small nucleotide polymorphisms with the risk of rectal cancer as well as the response to neoadjuvant chemoradiotherapy (nCRT) based on 5-Fluorouracil (5- FU)/leucovorin (LV) in the locally advanced setting.

**Patients and methods:** A total of 102 patients with locally advanced rectal cancer (LARC) and 119 healthy controls were included in this case-control study. Restriction fragment length polymorphism analysis (PCR-RFLP) was used for *MTHFR* genotyping

**Results:** Using dominant and recessive models, it was found that the *MTHFR* 667C allele and the 1298A allele were significantly associated with rectal cancer as low-penetrant factors. Combined genotype analysis highlighted the protective role of the 677CT/1298AC genotype and increased risk for rectal cancer development for carriers of 677CC/1298AA. Haplotype analysis indicated that carriers of haplotype 677C/1298A have an increased risk for rectal cancer development while the haplotype 677T/1298A has a protective role. No significant association with response to chemoradiotherapy was found

**Conclusion:** Our data point to *MTHFR* 667C allele and 1298A alleles as low-penetrant risk factors for rectal cancer in our population. To the best of our knowledge, this is the first study of this type performed on the Slavic population in the Western Balkan area which might be useful for future meta-analyses and the construction of genetic cancer risk prediction panels, as various population-based factors might also be significant in this setting.

## 1. Introduction

Colorectal cancer (CRC) is the third most frequently diagnosed cancer with 1.93 million newly diagnosed patients in 2020, and the second leading cause of cancer death worldwide with 930 000 deaths annually (Bray et al., 2018). The situation in Serbia is similar to the global one, CRC is the second most frequent cancer with regards to both incidence and mortality, with around 5000 newly diagnosed CRC cases and around 2400 deaths in 2020 (Serbian Cancer Registry, 2022). It is a multifactorial disease involving both genetic and environmental factors. Approximately 75% of CRCs are sporadic and occur in cases of absence of genetic predisposition or family history of CRC (Yamagishi et al., 2016). Diet plays an important role in the development of colorectal malignancy, as well as smoking history, alcohol consumption, body mass index (BMI), and lifestyle, although the relative significance of each of these factors on its own, or combinations of more than one factor is not clear (Ryan-Harshman and Aldoori, 2007). Rectal and colon cancer are different diseases with regard to diagnosis, sensitivity to treatment, and some risk factors, but few studies addressed risk factors for these two cancers separately.

The role of folic acid deficiency has been investigated in tumorigenesis in general (Kim, 2003). Some of the proposed models of folic depletion influence on cancer development are alterations in DNA methylation, disruption of DNA integrity, and disruption of DNA repair (Choi and Mason, 2000a). Interestingly, folate deficiency has been investigated as a factor only in alcohol-related carcinogenesis of rectal cancer, since there is a clear relationship between alcohol and alterations in folate metabolism (Choi and Mason, 2000b). Whether dietary intake of folate has a protective effect against selected cancers is not clear, since the results of studies are not consistent. An important gene in the metabolism of folic acid is the methylenetetrahydrofolate reductase (*MTHFR*) gene which provides instructions for making an enzyme called methylenetetrahydrofolate reductase (MTHFR). This enzyme converts the 5,10-methylenetetrahydrofolate acid to a different form of folate called 5- methyltetrahydrofolate which is the primary form of folate in blood. Two common functional polymorphisms in the *MTHFR* gene are C677T and A1298C. These polymorphisms influence enzyme activity. The C677T polymorphism in exon 4 causes a substitution of C to T nucleotide, leading to the substitution of alanine with valine in codon 222, which in turn affects the active site of the enzyme and thus reduces its activity, with the TT genotype product having a 70% reduced activity in comparison to wild type. A substitution of A to C at nucleotide 1298 (polymorphism A1298C of the *MTHFR* gene) leads to the substitution of glutamine with alanine, also causing reduced enzyme activity (Kennedy et al., 2012). The relationship between these polymorphisms and the risk of developing cancer has been researched and the results are contradictory and inconclusive. Some of the published studies showed differences in connection with ethnicity, as interestingly, there seems to be a higher frequency of the C677T TT genotype in southern Europe than in the north, while in Asia, the frequency is highest in China and lowest in India. Also, African Americans have a lower frequency of the TT genotype than Caucasians. The 1298CC genotype is more frequent in Caucasians (4-12%) than in China and Japan (1-4%) (Kennedy et al., 2012). The study conducted on patients with cervical cancer showed that *MTHFR* polymorphisms were associated with a higher risk of cervical cancer but not related to cervical intraepithelial neoplasia (CIN) (Gong et al., 2018). Zao et al. showed in their meta-analysis that the presence of the *MTHFR* C677T and A1298C polymorphisms is significantly associated with low risk of CRC (Zhao et al., 2013). A meta-analysis done on Asian population didn’t find a correlation between C677T polymorphism and the risk of developing CRC (Rai, 2015). A systematic review and meta-analysis of 67 studies suggested that *MTHFR* 677TT genotype is associated with a reduced risk of developing CRC only under conditions of high folate intake, but found no effect of A1298C polymorphism (Kennedy et al., 2012). Some studies also showed different influences of these polymorphisms on colon and rectal cancer. Komlosi et al. showed an effect of C677T on the risk of rectal but not colon cancer (Komlósi et al., 2010), Murtaugh et al. suggested a protective role against rectal cancer of *MTHFR* 677TT genotype in women, and 1298CC in men and women (Murtaugh et al., 2007), while a study in India showed no association of *MTHFR* 677T with colon cancer and a non-significantly increased risk of rectal cancer, and 1298CC genotype was associated with a significantly decreased risk of both rectal and colon cancer (Wang et al., 2006a). Study which aimed to evaluate the *MTHFR* gene polymorphisms in breast cancer patients, didn’t find an association with the risk of breast cancer development in general, but there was a relationship between both polymorphisms in relation to the risk of developing more aggressive biophenotypes of this cancer and, just in the case of A1298C there was an increased risk of lymph node metastasis (Castiglia et al., 2019). When the focus was on lung cancer, the results showed that C677T polymorphism is a risk factor and that A1298C gene polymorphism is a protective factor in lung cancer (Tong et al., 2018). All these results suggested that there are numerous factors influencing the effect of these polymorphisms on the risk of developing cancer.

Preoperative, neoadjuvant chemoradiotherapy (nCRT) based on 5-fluorouracil (5-FU), followed in most cases by operative treatment is the standard of care for LARC (Glynne-Jones et al., 2017). Tumor regression grade (TRG) is an established prognostic factor for local recurrence, disease-free, and overall survival (OS), with significantly better outcomes in patients showing TRG 1-2 (good responders) than those with TRG 3-5 (poor responders) (Vecchio et al., 2005). The cytotoxic activity of 5FU is mostly exhibited through an active metabolite which forms a complex with thymidylate synthase (TS) and 5,10- methylenetetrahydrofolate (5,10-MTHF) thus causing the inhibition of TS and disrupting normal DNA synthesis (Longley et al., 2003). Elevated intracellular MTHF levels are needed for optimal inhibition of TS, and these are controlled by methylene-tetrahydrofolate reductase (MTHFR), whose decreased activity results in higher levels of MTHF, higher formation of the 5,10-MTHF-TS complex and higher rate of inhibition of TS. There are large inter-individual differences in the efficacy of 5-FU and, considering the described mechanism of action of 5-FU, it is reasonable to assume that certain polymorphisms in genes involved in various points of 5-FU mechanism of action could explain some of these inter-individual differences in clinical response and toxicity to 5-FU (Ulrich et al., 2014). Polymorphisms in the *MTHFR* gene which cause decreased activity of MTHFR could make these patients more sensitive to 5-FU, therefore there should be a higher efficacy of 5-FU which in turn should lead to better survival. However, most study results do not prove this hypothesis. Two polymorphisms of the *MTHFR* gene, C677T and A1298C, have been investigated in this context in patients with CRC and more extensively in patients with LARC treated with chemo-irradiation. In this case, there is more consistency in the results of these studies with regard to the C677T polymorphism, with the C allele conferring a better response to nCRT in most studies, and a trend towards better DFS and OS in some. There doesn’t seem to be a major correlation between the A1298C polymorphism and efficacy of nCRT. In studies by Cecchin et al. (Cecchin et al., 2011) Garcia-Aguilar et al. (Garcia-Aguilar et al., 2011) and Ulrich et al. (Ulrich et al., 2014) the *MTHFR* 677T allele was significantly associated with a lower response, and in the study by Ulrich with a non-significant trend towards a worse DFS and OS in the homozygous state. In a study by Nikas et al. in 2015. It was shown that carriers of *MTHFR* 677CC genotype were 2.91 times more likely to have a good response to nCRT and 3.25 times less likely to have a relapse when compared to carriers of CT or TT genotypes (Nikas et al., 2015). Terrazzino et al. investigated *MTHFR* C677T and A1298C haplotypes and found that the haplotype *MTHFR* 677-T/1298-A was an independent predictor of tumor regression at univariate and multivariate analysis (Terrazzino et al., 2006). A meta-analysis by Salnikova and Kolobkov in 2015. on 45 papers involving CRT in LARC showed that *MTHFR* 677CC wild-type genotype was associated with a better response to nCRT compared with TT genotype (Salnikova and Kolobkov, 2016), and a systematic review by Zhao et al. suggested that there is a correlation between *MTHFR* C677T and tumor response using the recessive model in rectal cancer (Zhao et al., 2015). In contrast, Balboa et al. (Balboa et al., 2010) and Thomas et al. (Thomas et al., 2011) showed no association between these polymorphisms and response to chemo-irradiation in patients with rectal cancer. So far, various research which aimed to profile genetic risk factors of different types of cancer was conducted in Serbia in order to construct a general predictive risk model. (Cavic et al., 2019; Krivokuca et al., 2016; Cavic et al., 2016). Results like these might contribute to the construction of a low-cost and minimally invasive pan-cancer polymorphism screening tool.

In Serbia in the majority of cases, CRC is diagnosed in advanced stages where limited treatment options are available and survival is poor. Our group and others have invested efforts into profiling the diagnostic, prognostic and predictive factors for CRC and anal cancer, in an effort to provide better research strategies for treatment and overall management (Jakovljevic et al., 2012a; Brotto et al., 2013; Cavic et al., 2016; Nikolic et al., 2021; Stojanovic-Rundic et al., 2021; Vuletić et al., 2021; Marinkovic et al., 2023; Stanojevic et al., 2023).

The aim of this study was to analyze the association of *MTHFR* C677T and A1298C small nucleotide polymorphisms with the risk of rectal cancer as well as the response to neoadjuvant chemoradiotherapy based on 5-Fluorouracil (5-FU)/leucovorin (LV) in the locally advanced setting, in an effort to provide data from the Western Balkan area which is usually underrepresented in larger meta-analyses.

## 2. Material and Methods

### 2.1. In silico analysis using the Human Protein Atlas, UALCAN, ROCplotter, STRING, and NCBI GEOdatasets

The interactive web resource for analyzing cancer transcriptome data from the Cancer Genome Atlas (TCGA)(National Cancer Institute and the National Human Genome Research Institute, n.d.) database UALCAN (UALCAN database, n.d.; Chandrashekar et al., 2017) was used to analyze *MTHFR* expression levels in normal and rectal cancer samples. The publicly available database the Human Protein Atlas (HPA) (The Human Protein Atlas database V.20.0, n.d.; Uhlen et al., 2017) was used to analyze TCGA transcriptome data on the expression of *MTHFR* in relation to its prognostic significance in rectal cancer. Kaplan-Meier plots summarize the results of the correlation between *MTHFR* expression level and patient survival by dividing patients into low (under experimental cut-off) or high (above experimental cut-off) groups. Expression cut-off values for the HPA data are presented as the number fragments per kilobase of exon per million reads (FPKM) of *MTHFR* in the tumor tissue at diagnosis. ROCplotter (www.rocplot.org), an online tool that uses the transcriptome data of a large set of rectal cancer patients was used to analyze *MTHFR* expression levels in responders and non-responders (Fekete and Győrffy, 2019). Corresponding images and data were downloaded from the HPA, UALCAN and ROCplotter platforms in the original form. The STRING (STRING database, n.d.) protein network for MTHFR was built based on highest confidence (0.9) evidence from experimental and biochemical data, co-expression, gene neighborhood, gene co-occurrence, gene fusions, protein homology, manually curated metabolic and signaling pathway databases, and predictive and knowledge text data mining. The network included 5 primary-interaction shell proteins to explore interactions and clustering with other proteins and the effects of these interactions. For the enrichment analysis, the whole genome statistical background was assumed. The analysis was performed using STRING v.11.0 (Szklarczyk et al., 2019), corresponding images and results were exported and statistical significance was considered for p < 0.05. NCBI platform was used for the search of publicly available datasets for external validation of obtained results. Search criteria included the keyword “rectal cancer” while “genome variation profiling by SNP array” and “SNP genotyping by SNP array” were used as a study type of interest.

### 2.2. Patients and controls

A case-control study was performed in a group of 102 patients diagnosed with locally advanced primary rectal adenocarcinoma (age range 29-83 years, median 61; 68 males, 34 females) from several cancer centers in Serbia, and 119 healthy control subjects (age range 32-89 years, median 55; 67 males, 52 females) with no previous history of malignancies and no known folate metabolism deficiency, all of Caucasian descent (Table 1). All patients were diagnosed with locally advanced rectal cancer, stage II (T3/4N0M0) or III (T1-4N+M0) according to clinical and histological criteria of the 8th edition of the TNM classification of malignant tumors, and ECOG ≤ 2. (Oken et al., 1982) The tumors were located <15cm from the anocutaneous line and were treated with neoadjuvant chemoradiotherapy (5-Fluorouracil 350 mg/m2 i.v. bolus plus Leucovorin 25mg/m2 D1-D5 and D29-D33). Radiotherapy was initiated concurrently with chemotherapy, 50.4 Gy in 28 fractions, conventionally fractioned 1.8Gy/fr, using the technique with 3 or 4 radiation areas (all areas as recommended by the International Committee of Radiation Units and Measurements (ICRU) 50/62)(Landberg et al., 2016b, 2016a). Clinical response assessment took place 6-8 weeks after the completion of neoadjuvant therapy, involving pelvic MRI scans, rigid proctoscopy, and digital rectal examinations. Subsequently, patients were referred for surgery. The patohistological assessment of surgical specimens included the determination of histomorphology of the resected tumor (type and grade), tumor invasiveness (ypTNM, R classification)(Wei et al., 2018), pathohistological grading of the tumor regression by the Mandard scale(Siddiqui et al., 2016) with the determination of prognostic categories. Postoperative treatment was determined according to postoperative staging and response to neoadjuvant therapy. Follow-up of patients was conducted every three months during the first two years after completing treatment, and every six months thereafter. Clinical examinations and assessments of tumor markers (cancer embryonic antigen (CEA) and carbohydrate antigen (CA19-9)) were performed at each follow-up. CT/MRI scans of the abdomen and pelvis were scheduled every three months during the first year of follow-up and every six months thereafter. CT scan of thoranx and colonoscopies were conducted once a year.

**Table 1.** Patient characteristics.

| Characteristic | Patients<br>N (%) |
| --- | --- |
| **Age (years)** |  |
| Mean (SD) | 59.9 (9.9) |
| Median (Range) | 61.0 (38-76) |
| **Gender** |  |
| Female | 34 (33.3) |
| Male | 68 (66.7) |
| **UICC staging((UICC), 2020)** |  |
| II | 13 (12.8) |
| III | 89 (87.2) |
| **Grade** |  |
| 1 | 65 (63.8) |
| 2 | 31 (30.4) |
| 3 | 4 (4) |
| NA | 2 (2) |
| **Acute toxicity** |  |
| without | 22 (21.5) |
| grade I/II | 74 (72.5) |
| grade III/IV | 1 (1) |
| NA | 5 (5) |
| **RT technique** |  |
| 2D | 2 (2) |
| 3D | 100 (98) |
| **RT dose (Gy)** |  |
| Mean (SD) | 48.8 (2.9) |
| Median (Range) | 50.4 (36-50.4) |
| **Tumor response grade (TRG)\*** |  |
| 1 | 15 (14.7) |
| 2 | 15 (14.7) |
| 3 | 28 (28.4) |
| 4 | 35 (34.3) |
| 5 | 2 (2.9) |
| NA | 7 (7) |

\* According to Mandard scale(Siddiqui et al., 2016), NA - data unavailable

To investigate the predictive role of *MTHFR* polymorphisms, patients were divided into responders (patients with tumor regression grades (TRG) 1 and 2) and, non-responders (TRG 3-5) based on postoperative specimens. Patients who achieved a complete clinical response without subsequent operative treatments were excluded from this analysis.

### 2.3. DNA isolation and MTHFR genotyping

Genomic DNA was isolated from formalin-fixed paraffin-embedded tissue samples (FFPE) obtained by biopsy/resection using the QIAamp® DNA FFPE Tissue isolation kit (Qiagen, UK). Ethylenediaminetetraacetic acid (EDTA) peripheral blood was drawn from healthy controls by venipuncture and further used for leukocyte isolation using BloodPrep Chemistry for ABI PRISM™ 6100 Nucleic Acid PrepStation (Applied Biosystems, CA, USA). Recent large-scale literature analysis provided information that the concordance between germline and somatic DNA in variants of pharmacogenetic genes is virtually 100% [14]. The concentration and purity of the isolated DNA samples were determined spectrophotometrically (Nanodrop, Shimadzu). Restriction fragment length polymorphism analysis (PCR-RFLP) was used for *MTHFR* genotyping as previously described(Jakovljevic et al., 2012b; Cavic et al., 2014, 2016). The analysis was performed using the Agilent DNA 1000 Kit on the Agilent 2100 Bioanalyzer. To ensure adequate genotyping, a previously established heterozygote sample was used as a method of control, and genotyping was carried out blind to case-control status. Randomly selected 10% of samples were analyzed by Sanger sequencing to ensure data validity.

### 2.4. Statistical analyses

Descriptive methods of statistical analysis (frequencies, percentage, mean, median, standard deviation /SD/ and range) were used to summarize the sample data. The Hardy-Weinberg equilibrium of the analyzed polymorphisms was tested using the Pearson Chi-Square test. Two-sided p values <0.05 were considered to indicate statistical significance. The associations between the patients’ and healthy controls as well as responders and non responder were analyzed using Pearson Chi-Square with Yates’ correction. Fisher’s exact test was used for analyzing differences between males and females. Combined genotype frequencies were calculated by direct counting while statistical significance in combined genotype distribution between patients and controls were observed using Chi-Square test with Yates’ correction. In addition, haplotype analysis was used for calculating the interaction between two polymorphic sites of *MTHFR*. Haplotype frequencies were calculated manually, and data were confirmed using Multiallelic Interallelic Disequilibrium Analysis Software (University of Southampton, Highfield, Southampton, UK) (Gaunt et al., 2006) and Golden Helix Tree SNP & Variation Suite software. Statistical significance was obtained using Pearson Chi-Square with Yates’ correction. All statistical analysis was performed using GraphPad Prism 8.0.1.

## 3. Results

### 3.1. In silico analyses

*In silico* analyses using the UALCAN and HPA platforms showed that *MTHFR* expression was significantly different higher in normal compared to rectal cancer tissue (Fig1a, p<0.01) and that its low expression is correlated with higher cancer stages (Fig1b; stage 1 vs. stage 3, p<0.02; stage 1 vs. stage 4, p<0.05). It was generally not prognostically significant in rectal cancer (p=0.27), but the 5-year survival rate was found to be 91% for the high expression group and 48% for the low expression group (Fig1c; expression cut-off 3.14 FPKM, median follow up time 1.75 years).

**Fig 1.**
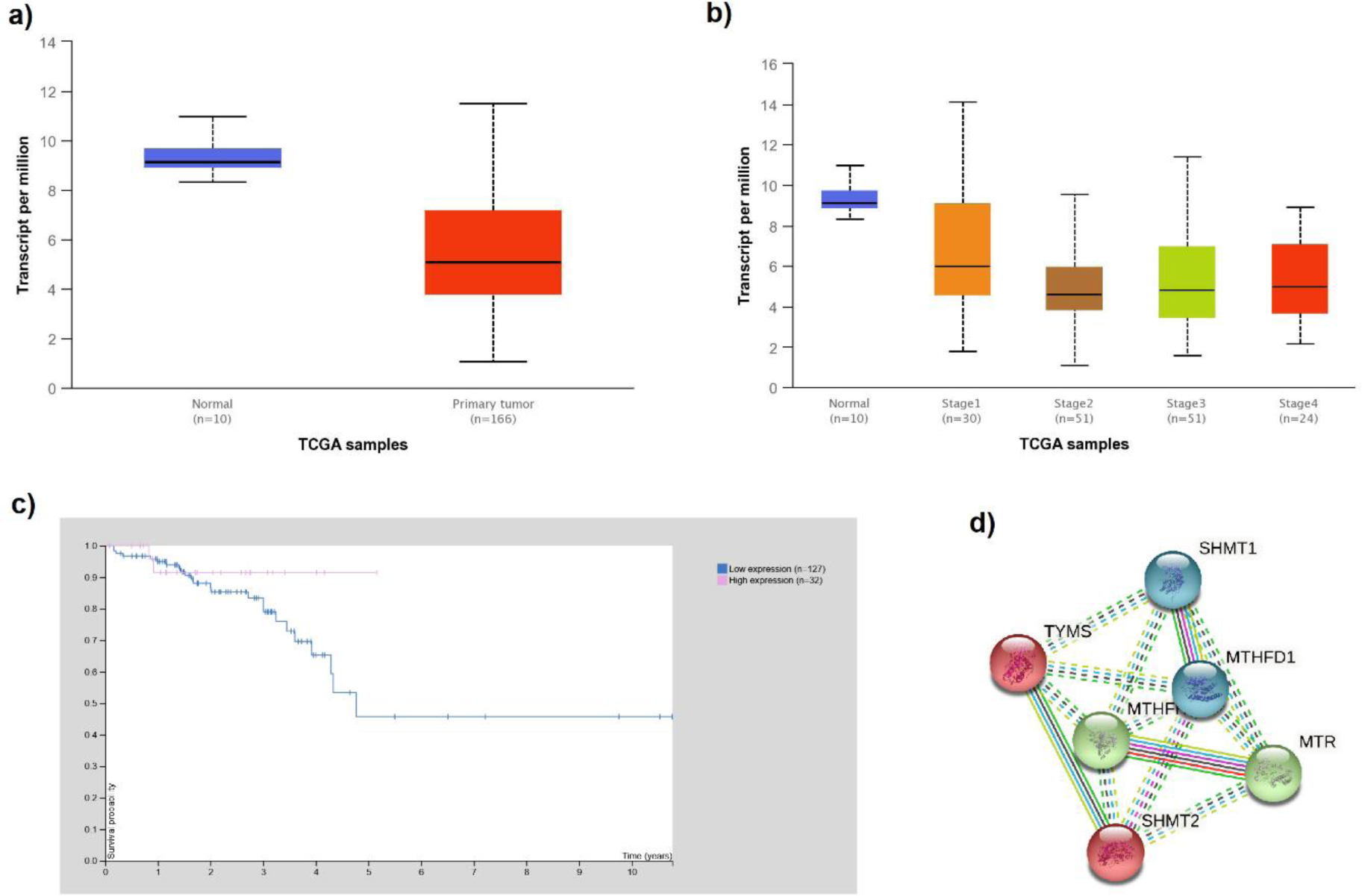
Expression of *MTHFR* in rectal cancer according to the UALCAN and HPA analysis of TCGA data. a) Expression of *MTHFR* in rectal adenocarcinoma based on sample type (normal vs. cancer tissue p<0.01). b) Expression of *MTHFR* in rectal adenocarcinoma based on individual cancer stage (stage 1 vs. stage 3, p<0.02; stage 1 vs. stage 4, p<0.05). c) Survival curves of rectal cancer patients in relation to the expression of *MTHFR* (expression cut-off 3.14 FPKM, p=0.27). c) Direct STRING (STRING database, n.d.) network of MTHFR was built based on highest confidence (0.9) evidence from experimental interaction data (pink), co-expression (black), gene neighborhood (green) and co-occurrence (blue) data, curated databases (light blue), predictive and knowledge text mining (light green), protein homology (purple). The network included 5 primary-interaction shell genes to explore their indirect interactions and clustering on all analyzed platforms (PPI enrichment p-value: 0.002). Red nodes – TYMS cluster members; green nodes – MTHFR cluster members; blue nodes – MTHFD1 cluster. Nodes are labeled with HGNC symbols: MTHFD1 - C-1- tetrahydrofolate synthase; MTHFR - Methylenetetrahydrofolate reductase; MTR - Methionine synthase; TYMS - Thymidylate synthase; SHMT1 - Serine hydroxymethyltransferase, cytosolic; SHMT2 - Serine hydroxymethyltransferase, mitochondrial.

ROCplotter analysis included 42 patients in total (19 responders and 23 non-responders) and highlighted that *MTHFR* expression (Affy ID 7436) is slightly higher in a group of patients who responded poorly to the therapy but without statistical significance. There was weak statistical significance in ROC p-value (p=0.045) with AUC=0.648.

The STRING analysis showed that *MTHFR* has reported direct and indirect interactions with various proteins that are important for rectal cancerogenesis/homeostasis when gene co-expression, experimental/biochemical data, and text mining were considered at the highest confidence level (0.9) (Fig.1d). Cluster analysis extended to 5 primary-interaction shell genes (MTHFD1 - C-1-tetrahydrofolate synthase, MTR - Methionine synthase, TYMS - Thymidylate synthase, SHMT1 - Serine hydroxymethyltransferase, cytosolic and SHMT2 - Serine hydroxymethyltransferase, mitochondrial) showed that these proteins form biological clusters across all analyzed platforms. The extended network was found to be enriched in interactions (PPI enrichment p-value: 0.002), which indicated that they *i*nteract with each other significantly more than is expected for a random set of proteins of similar size and can be thus considered as a biologically interconnected group (Szklarczyk et al., 2019).

### 3.2. MTHFR genotyping

Patient characteristics are presented in Table 1. Of the 102 analyzed patients, *MTHFR* genotyping was successful in 97 patients (95%) and in all 119 healthy controls. The quality of DNA isolated from the remaining 5 FFPE samples was too low for successful genotyping. A 198 bp PCR product containing the *MTHFR* C677T polymorphic site was successfully obtained from all patient and control samples (Fig.2a). After digestion of the PCR products, an undigested PCR product (198 bp) indicated the presence of a homozygous wild-type genotype (CC), while heterozygotes (CT) produced three bands of 198, 175 and 23 bp, and homozygotes (TT) produced two fragments of 175 and 23 bp. A 163 bp PCR product containing the *MTHFR* A1298C polymorphic site was successfully obtained from all patient and control samples (Fig.2b). After digestion of the PCR products, the presence of a homozygous wild type A allele leads to the appearance of 5 bands of 56, 31, 30, 28 and 18 bp, while the presence of the C allele leads to the appearance of 4 bands of 84, 31, 30 and 18 bp.

**Fig 2.**
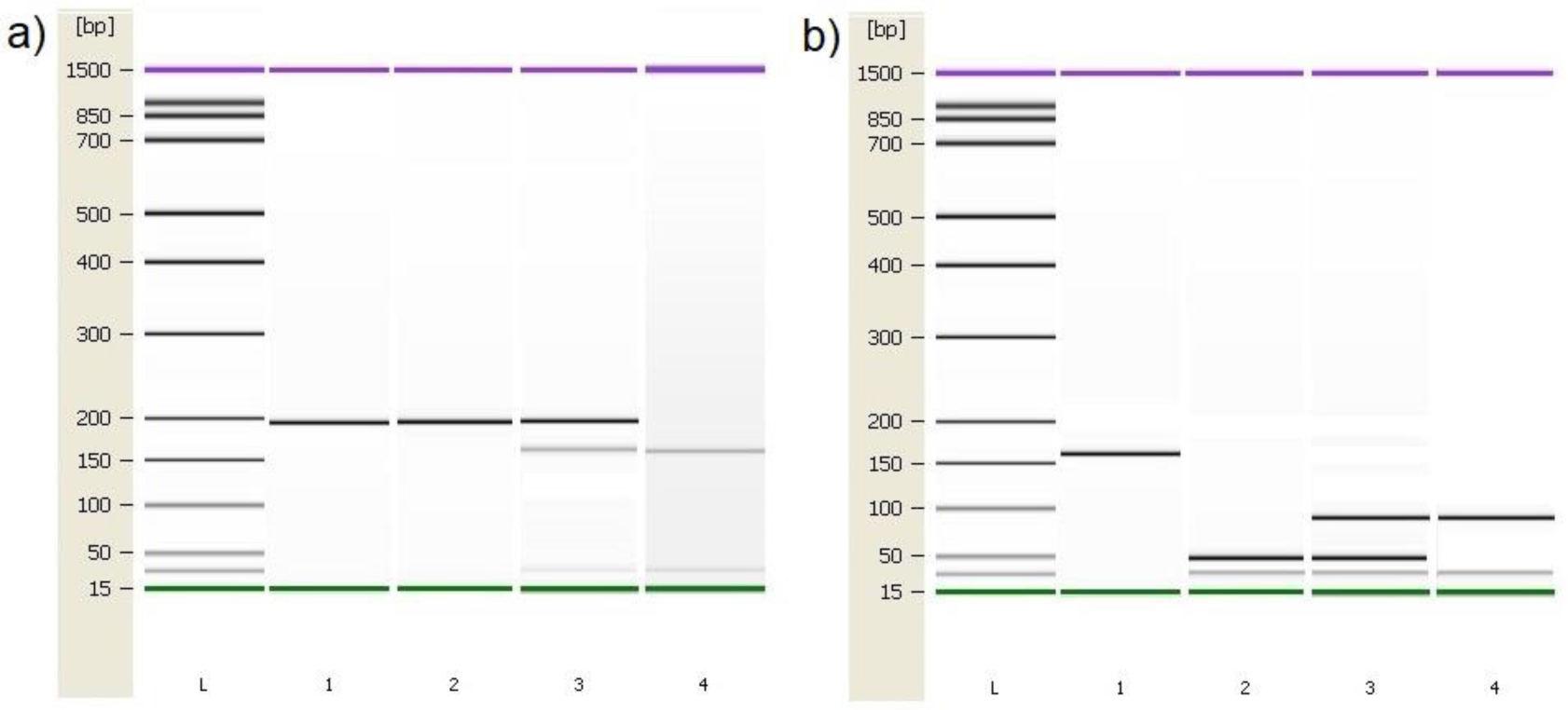
Genotyping results of the *MTHFR* C677T and A1298C polymorphic variants using the Agilent 2100 Bioanalyzer. a) PCR and RFLP results of the *MTHFR* C677T polymorphic variant. Column 1: 198 bp PCR product. Column 2: C/C, Column 3: C/T, Column 4: T/T. b) PCR and RFLP genotyping results of the *MTHFR* A1298C polymorphic variant. Column 1: 163 bp PCR product. Column 2: A/A, Column 3: A/C, Column 4: C/C. L – High-sensitivity DNA ladder (Agilent Technologies). 1500 bp upper and 15 bp lower marker are present in each column.

### 3.3. Significance for cancer risk

The distribution of *MTHFR* C677T genotypes in patients and controls did not deviate from the Hardy-Weinberg equilibrium (Table 2, χ2=3.34; p=0.069 and χ2=3.59; p=0.058). The allele frequencies of the *MTHFR* C677T polymorphic variants in patients and controls showed that the frequency of the C allele was higher in patients (0.74) than in healthy controls (0.60) (Fig3.a-b). CC homozygosity at the 677 polymorphic site of the *MTFHR* gene was more prevalent in patients with rectal cancer than in healthy control subjects. The *MTHFR* C allele was associated with rectal cancer in both the dominant and the recessive models (Table 3). In this study, the TT genotype seems to have a protective effect on the development of rectal cancer in males (p=0.0304), while no significant correlation was found in female patients.

**Table 2.**
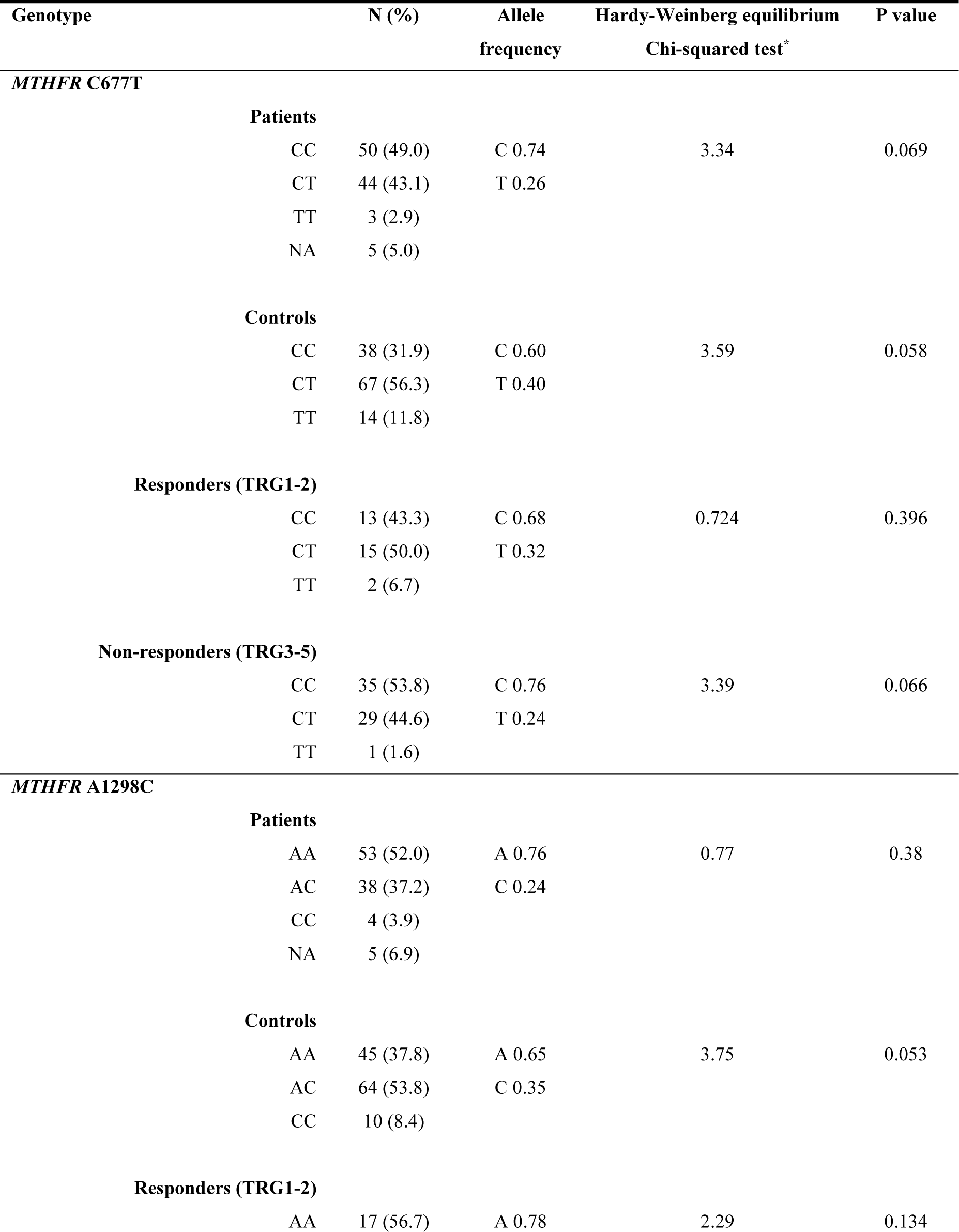
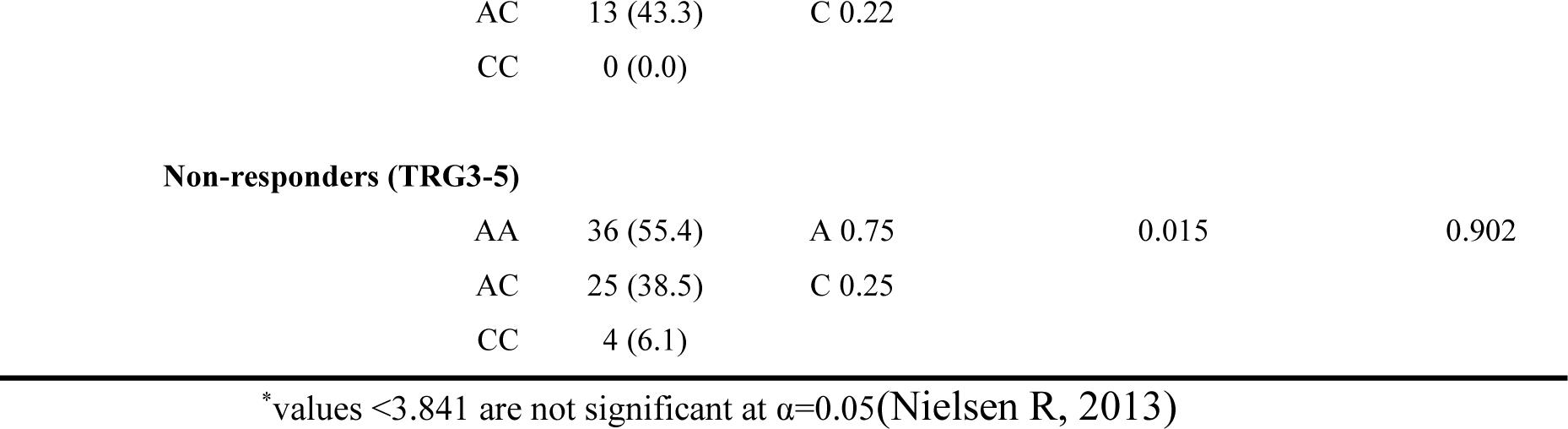
Genotype and allele frequencies of *MTHFR* C677T and A1298C polymorphic variants in the patient and healthy control groups.

**Table 3.** Analysis of the effects of *MTHFR* C677T and A1298C polymorphic variants on rectal cancer risk using dominant and recessive models.

| Gene / Model | Genotype | Patients vs. Healthy controls<br>OR (95% CI) | p value<br>(Pearson $\chi^2$ test) |
| --- | --- | --- | --- |
| *C677T* |  |  |  |
| *dominant model* | CC vs. any T | 2.37 (1.39 - 4.03) | **0.002** |
| *recessive model* | Any C vs. TT | 3.67 (1.17 - 11.50) | **0.0345** |
| *A1298C* |  |  |  |
| *dominant model* | AA vs. any C | 2.0 (1.20 - 3.59) | **0.013** |
| *recessive model* | any A vs. CC | 2.08 (0.63 - 6.88) | 0.340 |

statistically significant results are labeled bold; <sup>1</sup>statistical trend.

**Table 4.** Analysis of the effects of *MTHFR* C677T and A1298C polymorphic variants on response to neoadjuvant chemoradiotherapy using dominant and recessive models.

| Gene / Model | Genotype | Good vs. poor<br>responders<br>OR (95% CI) | p value<br>(Pearson $\chi^2$ test) |
| --- | --- | --- | --- |
| *C677T* |  |  |  |
| *dominant model* | CC vs. any T | 0.65 (0.27 - 1.57) | 0.464 |
| *recessive model* | Any C vs. TT | 0.22 (0.02 - 2.51) | 0.4855 |
| *A1298C* |  |  |  |
| *dominant model* | AA vs. any C | 1.05 (0.44 - 2.52) | 0.907 |
| *recessive model* | any A vs. CC | n.d. | 0.406 |

**Fig 3.**
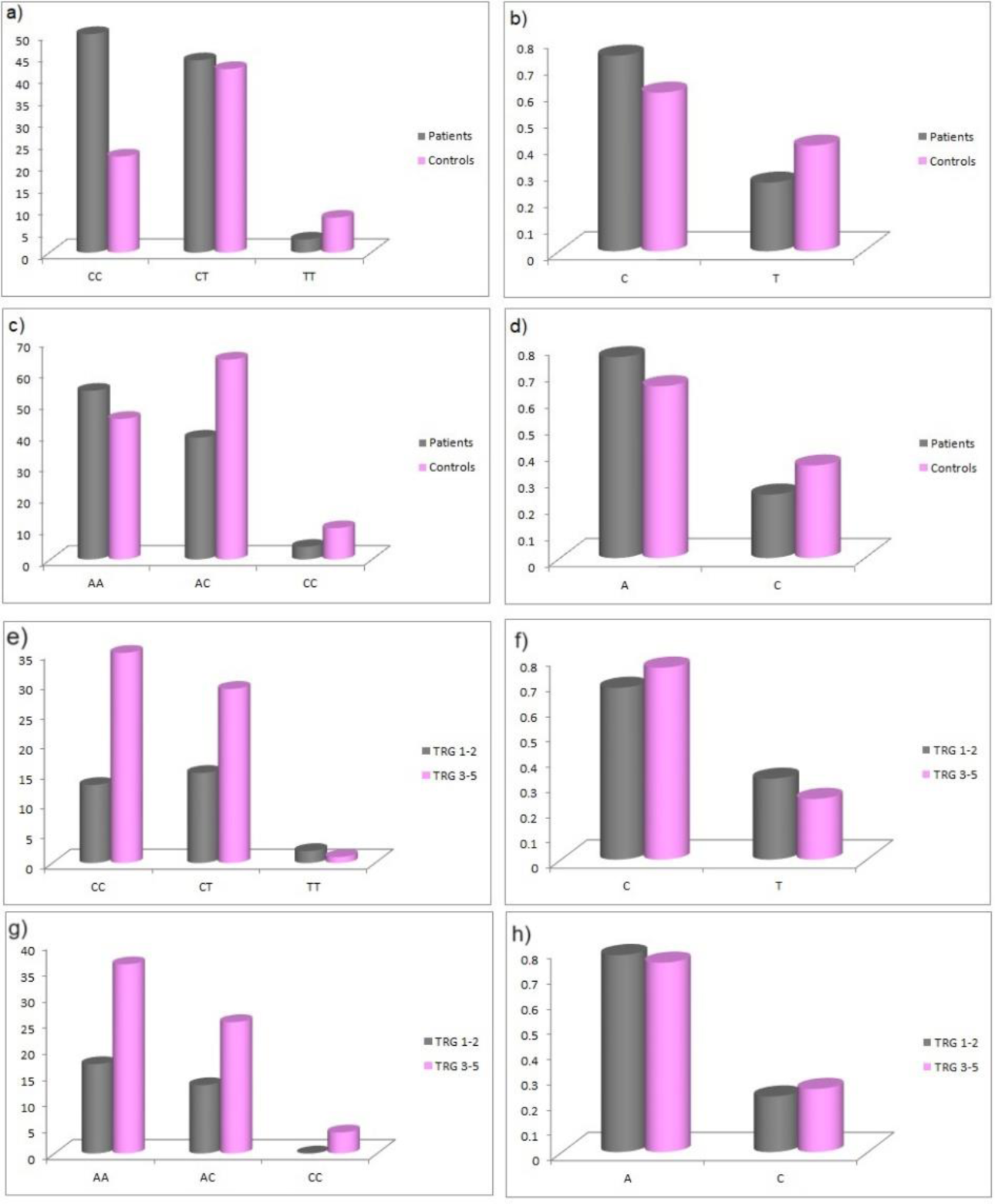
a) Genotype and b) allele distribution of the *MTHFR* C677T polymorphic variants in rectal cancer patients and healthy controls. c) Genotype and d) allele distribution of the *MTHFR* A1298C polymorphic variants in rectal cancer patients and healthy controls. e) Genotype and f) allele distribution of the *MTHFR* C677T polymorphic variants in responders and non-responders to neoadjuvant chemoradiotherapy. g) Genotype and h) allele distribution of the *MTHFR* A1298C polymorphic variants in responders and non-responders to neoadjuvant chemoradiotherapy.

The distribution of *MTHFR* A1298C genotypes in patients and controls did not deviate from the Hardy-Weinberg equilibrium (Table 1, χ2=2.17; p=0.0141 and χ2=3.78; p=0.051). Frequency of the C allele was slightly higher in patients (0.76) than in healthy controls (0.65) (Fig3.c-d). AA homozygosity at 1298 polymorphic site of the *MTFHR* gene was associated with higher risk for developing rectal cancer in the dominant model (Table 3). There were no differences according to sex with regards to the effect of this polymorphism and risk for rectal cancer.

### 3.4. Combined genotype

In groups of patients and controls, the 4 common combined genotypes were CT/AA (29.90% vs. 19.33%), CC/AC (24.74% vs. 15.97%), CC/AA (24.74% vs. 12.61%), and CT/AC (13.40% vs. 32.77%) (Table 5). Statistical analysis of combined genotypes highlighted the protective role of CT/AC combined genotype (p=0.0016) while CC/AA genotype showed an increased risk for rectal cancer development (p=0.0162) (Table 6).

**Table 5.** Combined genotype frequencies of *MTHFR* C677T and A1298C polymorphic variants within the patients and controls.

| *Combined*<br>*genotype* | N (%)<br>Patients | N (%)<br>R | N(%)<br>NR | N (%)<br>Controls |
| --- | --- | --- | --- | --- |
| **CT/AA** | **29 (29.9)** | **10 (33.3)** | **19 (29.2)** | **23 (19.3)** |
| **CC/AC** | **24 (24.7)** | **6 (20.0)** | **17 (26.1)** | **19 (16)** |
| **CC/AA** | **24 (24.7)** | **7 (23.3)** | **16 (24.6)** | **15 (12.6)** |
| **CT/AC** | **13 (13.4)** | **5 (16.7)** | **8 (12.3)** | **39 (32.8)** |
| TT/AC | 2 (2.1) | 2 (6.7) | 0 (0) | 6 (5.0) |
| CT/CC | 2 (2.1) | 0 (0) | 2 (3.1) | 5 (4.2) |
| CC/CC | 2 (2.1) | 0 (0) | 2 (3.1) | 4 (3.4) |
| TT/AA | 1 (1.0) | 0 (0) | 1 (1.5) | 7 (5.9) |
| TT/CC | 0 (0.0) | 0 (0) | 0 (0) | 1 (0.8) |

Combined genotypes with the highest prevalence are labeled bold

**Table 6.** The effects of *MTHFR* C677T and A1298C combined genotype on rectal cancer risk.

| Combined genotype | Patients vs. Healthy controls OR (95% CI) | p value (Pearson $\chi^2$ test) |
| --- | --- | --- |
| CC/AA vs any other | 2.499 (1.246-5.081) | **0.0162** |
| CT/AC vs any other | 3.15 (1.576-6.279) | **0.0016** |

statistically significant results are labeled bold

### 3.5. Haplotype analysis

Haplotype analysis indicated that the most frequent haplotypes in patients vs. controls were CA (677C-1298A) (46.15% vs. 32.99%) and TA (677T-1298A) (24.10% vs. 26.04%) followed by CC (677C-1298C) (21.03% vs. 23.26%). The rarest haplotype was TC (677T- 1298C) (8.72% vs. 17.71%). The carriers of the CA haplotype had the highest risk for developing rectal cancer (OR: 1.74; 95%CI 1.198-2.530, p=0.002) while the TC haplotype seems to provide a protective effect. (OR: 0.44; 95%CI 0.248-0.795, p=0.003). These results indicate that the two loci 677 and 1298 share relatively weak linkage disequilibrium in the patient group (D’= 0.27, r^2^= 0.00797) (Table 7).

**Table 7.** Estimated haplotype frequencies of *MTHFR* C677T and A1298C polymorphisms in patients and controls.

| Haplotype | N (%)<br>patients | N (%)<br>controls | OR (95% CI) | p value |
| --- | --- | --- | --- | --- |
| 677C-1298A | 90 (46.15) | 95 (32.99) | 1.74 (1.190-2.510) | **0.005** |
| 677C-1298C | 41 (21.03) | 67 (23.26) | *ns* | *ns* |
| 677T-1298A | 47 (24.10) | 75 (26.04) | *ns* | *ns* |
| 677T-1298C | 17 (8.27) | 51 (17.71) | 2.25 (1.286-3.977) | **0.008** |

statistically significant results are labeled bold

### 3.6. Significance for response to chemoradiotherapy

The distribution of *MTHFR* C677T genotypes in responders and non-responders did not deviate from the Hardy-Weinberg equilibrium (Table 2, χ2=0.724; p=0.396 and χ2=3.39; p=0.066). The allele frequencies of the *MTHFR* C677T polymorphic variants in responders and non-responders showed that the frequency of the C allele was lower in responders (0.68) than in non-responders (0.76) (Fig3.e-f) but no statistical significance was obtained.

The distribution of *MTHFR* A1298C genotypes in responders and non-responders did not deviate from the Hardy-Weinberg equilibrium (Table 2, χ2=2.29; p=0.134 and χ2=0.152; p=0.902). The frequency of the A allele was lower in responders (0.68) than in non-responders (0.76) (Fig3.g-h) but no statistical significance in predictive potential was obtained. Within the groups of responders and non-responders no statistical significance was obtained in terms of predictive potential of combined genotypes as well as haplotype frequencies. Haplotype analysis indicated that the two loci 677 and 1298 show relatively strong linkage disequilibrium in the non-responder group (D’= 0.46, r^2^= 0.02163).

### 3.7. Validation of results

Seven studies were found, but after initial processing one study, GSE35282 met all criteria and included polymorphic variant A1298C data of *MTHFR* from 43 patients with diagnosed locally advanced rectal cancer. There was no study available with C677T polymorphic variant of *MTHFR*. (Kim et al., 2013) Patients were divided according to Mandard TRG status into responders (TRG1/2; 41.9%) and non-responders (TR3/4; 58.1%, there were no patients with TRG5). Data obtained in mentioned study shown high correlation with our results. Statistical significance was not found within the predictive potential of the A1298C *MTHFR* polymorphic variant while allele frequencies showed a high correlation between these two studies (Table 8).

**Table 8.** External validation of obtained results using publicly available dataset GSE35282.

| A1298C polymorphic site<br>*MTHFR* | Allele Frequency<br>GSE35282 | Allele Frequency<br>our results | p value<br>Pearson $\chi^2$ test |
| --- | --- | --- | --- |
| **Patients** | A 0.84 | A 0.76 | ns |
|  | C 0.13 | C 0.24 |  |
| **Responders** | A 0.77 | A 0.78 |  |
|  | C 0.23 | C 0.22 |  |
| **Non-responders** | A 0.84 | A 0.76 |  |
|  | C 0.16 | C 0.24 |  |

## 4. Discussion

Since colorectal cancer remains one of the most common and deadly malignant diseases worldwide, much effort is directed to elucidating the etiopathological mechanisms and risk factors. Some risk factors are well known such as diet, alcohol consumption, smoking history and lifestyle but they alone are not enough to explain the development of the disease in all patients. Folate metabolism, more precisely folate deficiency is a very well-known factor of tumorigenesis in general since it plays an important role in maintaining genomic stability by being involved in DNA synthesis, repair, and methylation (Levin and Varga, 2016). One of the central enzymes involved in folate metabolism is 5,10-methylenetetrahydrofolate reductase (MTHFR) which irreversibly converts 5,10-methylenetetrahydrofolate to 5- methylenetetrahydrofolate that provides a methyl group for synthesis of methionine, whose deficiency may affect DNA synthesis (Ryan and Weir, 2001). Two common functional polymorphisms in the *MTHFR* gene that have been found to have an influence on the risk of CRC are C677T and A1298C. These SNPs cause reduced activity of the MTHFR enzyme, thus influencing folate metabolism, causing folate depletion and DNA hypomethylation, and disruption of DNA synthesis and repair (Kennedy et al., 2012). DNA hypomethylation is a nearly universal early event in carcinogenesis. It has been suggested that site-specific DNA hypomethylation may be critical, such as the hypomethylation of the coding region (exons 5-8) of the p53 tumor suppressor gene, which is the most frequently mutated in human cancers (Kim et al., 1997; Choi and Mason, 2000b).

Data regarding the specific influence of these polymorphisms on CRC risk are varied and inconclusive, and importantly, very few studies investigated the effect of these polymorphisms on the risk of rectal and colon cancer separately, which could be different since rectal and colon cancer are different diseases in their pathogenesis, histology, and sensitivity to treatment. We aimed to investigate the effect of C677T and A1298C polymorphisms on the risk of developing rectal cancer only, in the population of patients in Serbia. The results of this case-control study on 102 patients diagnosed with LARC and 119 healthy volunteers showed a higher frequency of the C677T C allele in patients than in controls (0.74), and the CC homozygosity at the 677 polymorphic sites was more common in patients than in healthy controls. This would suggest a protective effect of the T allele against rectal cancer. These results were confirmed using both the dominant and the recessive model of association. Data from previous studies are inconclusive, with most studies being carried out on small, ethnically homogenous populations, as is ours. Murtaugh et al. in United States (Murtaugh et al., 2007), Levine et al. in Canada (Levin and Varga, 2016), Sheng et al. (Sheng et al., 2012), and Rai et al (Rai, 2016) in Asia showed the protective effect of the 677 T allele against CRC and rectal cancer in the American study, as in the situation in our study. In a meta-analysis of 67 studies carried out in 25 countries over the world and comprising all ethnic groups, it was found that the homozygous variant genotype *MTHFR* 677TT confers a reduced risk of CRC by 12%, but the risk between carriers of 677CT and CC genotypes was similar (Kennedy et al., 2012). On the other hand, a meta-analysis by Teng and colleagues in 2013. done on 71 studies and over 30 thousand patients showed an increased risk of CRC in carriers of the 677TT genotype, but only in Caucasian patients and not Asians (Teng et al., 2013). The same results were found in the Indian and the Hungarian population, with the caveat that this effect was found only on patients with rectal cancer in Hungary, but not colon cancer (Wang et al., 2006b; Komlósi et al., 2010). A recent paper by Alanov and colleagues in Azerbaijan showed no effect of this polymorphism on the risk of CRC (Aslanov et al., 2023).

In our group, there was a slightly higher frequency of *MTHFR* 1298C allele (0.75) than in controls (0.65), and AA homozygosity at the 1298 polymorphic site was more common in patients than in healthy controls. As with the C677T polymorphism, data about the effect of A1298C polymorphism are not consistent. In a study by Jiang and colleagues, carriers of the 1298C allele had a lower risk of rectal cancer (Jiang et al., 2005). Same association was found in the Indian study where carriers of the 1298AC genotype were at a lower risk for colon cancer (OR 0.43, 95% CI 0.22-0.82) and rectal cancer (OR 0.7), and carriers of the homozygous CC genotype had a significantly lower risk for both colon (OR 0.3, 95% CI 0.09-0.80) and rectal cancer (OR 0.43, 95% CI 0.23-0.80) (Wang et al., 2006b). This data would suggest a protective effect of the C allele of the A1298C polymorphisms. Again, a 33% reduction in the risk of rectal cancer in individuals carrying the 1298CC genotype was reported (Murtaugh et al., 2007). On the other hand, a Japanese case-control study on 220 patients with rectal cancer and controls found no association between this polymorphism and risk of rectal cancer nor did the meta-analysis of Kennedy and colleagues (Matsuo et al., 2005). However, in this meta-analysis, there was a lower risk of CRC in carriers of the 1298CC genotype in Asian and American studies, but an increased risk of CRC in European countries, which would suggest some geographic differences, possibly related to dietary habits of folate intake or alcohol consumption.

Very few studies stratified results also based on sex, but as before, results are inconclusive and inconsistent. Lightfoot et al found a reduced risk of CRC in men with the 677CT genotype, and an increased risk in women with the 677TT genotype, while most other studies didn’t find any difference (Lightfoot et al., 2008). Komlosi et al. investigated the Hungarian population and stratified results according to age and sex, finding that the presence of the 677 C allele increased the risk for rectal cancer only in younger (<60) and male patients, but found no influence on colon cancer risk (Komlósi et al., 2010). On the other hand, Murtaugh et al. found a reduced risk of rectal cancer only in female carriers of the 677 T allele, especially in women 60 years or older (OR 0.32, 95% CI) in the United States (Murtaugh et al., 2007). Our analysis highlighted the protective role of the 677TT genotype for rectal cancer development only in males (p=0.0304).

There is not much data on the combined effect of *MTHFR* C677T and A1298C polymorphisms and risk of CRC. It has been previously described that the *MTHFR C677T* and *A1298C* polymorphisms are in linkage disequilibrium, which means that combinations of *677CT* and *1298CC*, *677TT* and *1298AC*, and *677TT* and *1298CC* are very rare (Kono and Chen, 2005). In this review, it was found that having one variant allele of either *677T* or *1298C* does not affect the risk of colorectal cancer. In the study by Murtaugh et al the lowest risk of CRC was found in carriers of 677CC/1298CC. However, in the meta-analysis by Kennedy and the study of Yin et al carriers of the 677TT/1298AA had the lowest risk of CRC, and carriers of 677CC/1298CC had a non-significantly higher risk of CRC in Yin’s study, again, showing inconsistencies in literature data (Yin et al., 2004). Our results suggest that individuals with haplotypes 677C and 1298A have an increased risk for rectal cancer development compared to any other haplotype while 677T and 1298C haplotypes have a protective role. Combined genotype analysis highlighted that individuals with CC/AA genotype combination have an increased risk for rectal cancer development while CT/AC genotype has a protective role.

Although our analysis did not show a statistically significant association between *MTHFR* C677T and A1298C polymorphisms and response to therapy, our results are somewhat different than most previously reported in the literature. We found a slightly lower frequency of the 677C allele in responders (0.68) than in non-responders (0.76), suggesting that carriers of C allele are less likely to respond to nCRT. Most other studies (Longley et al., 2003; Vecchio et al., 2005; Balboa et al., 2010; Cecchin et al., 2011; Garcia-Aguilar et al., 2011; Thomas et al., 2011; Ulrich et al., 2014; Zhao et al., 2015; Salnikova and Kolobkov, 2016) found a relationship with the 677C allele and better response to nCRT, which goes further to suggest a very complex and probably multifactorial influence of genetic and other factors on the efficacy of nCRT. The most frequent explanation of the effect of *MTHFR* polymorphisms on the efficacy of 5FU-based nCRT is that the decreased activity of MTHFR causes elevated levels of 5,10-MTHF, thus more formation of the complex with TS reducing TS activity and disrupting DNA and RNA synthesis (Longley et al., 2003; De Mattia et al., 2020).

However, another hypothesis focuses on the higher availability of non-methylated folate substrates in patients with reduced MTHFR activity, which can be used for de novo synthesis of DNA, thus preserving DNA integrity. Also, carriers of the 677TT genotype could be less prone to DNA damage by radiotherapy, thus making the nCRT less effective in these patients (Kawakami et al., 2003; Leopardi, 2006).

All this emphasizes the complexity and multifactorial influences on treatment outcomes, especially in patients treated with combined modalities.

## 5. Conclusions

Data obtained in this study point to low-cost, non-invasive, and easily determined factors that might be helpful to identify specific subgroups of patients that should be monitored more closely, which is especially important in developing countries. The *MTHFR* 667C allele and 1298A alleles were identified as low-penetrant risk factors for rectal cancer in our population. Our study did not show the influence of these polymorphisms on the efficacy of nCRT, but a further and more detailed analysis using a wider panel of genetic factors is planned. To the best of our knowledge, this is the first study on this subject performed on the Slavic population in the Western Balkan area which might be useful for future meta-analyses, as various population-based factors might also be significant in this setting.

## Acknowledgements

This study was funded by the Horizon Europe Twinning Project STEPUPIORS (Agreement No. 101079217) and the Ministry of Education and Science of the Republic of Serbia (Agreement No. 451-03-47/2023-01/200043).

## Author contributions

Conceptualization (AS, JS, MM, MC); Data curation (AS, JS, MM, MC); Formal analysis (AS, JS, MM); Funding acquisition (MC, RF, SCB, JZ); Investigation (AS, JS, SSR, RJ); Methodology (AS, MM, MC); Project administration (AD, MC, RJ, JZ, RF, SCB); Resources (SSR, RJ, AD, MC, RJ, JZ, RF, SCB); Software (AS, MM); Supervision (SSR, RJ, MC, RJ, JZ, RF, SCB); Validation (AS, JS, AD); Writing—original draft (AS, JS, MM, MC); Writing—review and editing (AS, JS, MM, SSR, RJ, AD, JZ, RF, SCB, MC). All authors contributed to the article and approved the submitted version.

## Study approval

The procedures used in this study were approved by the Ethics Board of the Institute for Oncology and Radiology of Serbia and were in accordance with the Helsinki Declaration of 1964 and its later amendments or comparable ethical standards. All patients signed an informed consent.

## Data Availability Statement

The data that support the findings of this study are available upon reasonable request from the corresponding author. The data are not publicly available due to ethics restrictions (their containing information could compromise the privacy of patients).

## References

1. Aslanov, H., Agaev, R., Bayramov, B., Hadizade, A., Alakbarova, N., Abdulrahimli, S., et al. (2023). Research Article Lack of significant association between MTHFR gene C677T polymorphism and colorectal cancer in the Azerbaijani population. Genetics and Molecular Research 22. doi: 10.4238/gmr19100.

2. Balboa, E., Duran, G., Lamas, M. J., Gomez-Caamaño, A., Celeiro-Muñoz, C., Lopez, R., et al. (2010). Pharmacogenetic analysis in neoadjuvant chemoradiation for rectal cancer: high incidence of somatic mutations and their relation with response. Pharmacogenomics 11, 747–761. doi: 10.2217/pgs.10.51.

3. Bray, F., Ferlay, J., Soerjomataram, I., Siegel, R. L., Torre, L. A., and Jemal, A. (2018). Global cancer statistics 2018: GLOBOCAN estimates of incidence and mortality worldwide for 36 cancers in 185 countries. CA Cancer J Clin 68, 394–424. doi: 10.3322/caac.21492.

4. Brotto, K., Malisic, E., Cavic, M., Krivokuca, A., and Jankovic, R. (2013). The Usability of Allele-Specific PCR and Reverse-Hybridization Assays for KRAS Genotyping in Serbian Colorectal Cancer Patients. Dig Dis Sci 58, 998–1003. doi: 10.1007/s10620-012-2469-9.

5. Castiglia, P., Sanna, V., Azara, A., De Miglio, M. R., Murgia, L., Pira, G., et al. (2019). Methylenetetrahydrofolate reductase (MTHFR) C677T and A1298C polymorphisms in breast cancer: a Sardinian preliminary case-control study. Int J Med Sci 16, 1089– 1095. doi: 10.7150/ijms.32162.

6. Cavic, M., Krivokuca, A., Boljevic, I., Brotto, K., Jovanovic, K., Tanic, M., et al. (2016). Pharmacogenetics in cancer therapy - 8 years of experience at the Institute for Oncology and Radiology of Serbia. J BUON 21, 1287–1295.

7. Cavic, M., Krivokuca, A., Spasic, J., Brotto, K., Malisic, E., Radulovic, S., et al. (2014). The influence of methylenetetrahydrofolate reductase and thymidylate synthetase gene polymorphisms on lung adenocarcinoma occurrence. J BUON 19, 1024–1028.

8. Marinkovic, M., Stojanovic-Rundic, S., Stanojevic, A., Ostojic, M., Gavrilovic, D., Jankovic, R., et al. (2023). Exploring novel genetic and hematological predictors of response to neoadjuvant chemoradiotherapy in locally advanced rectal cancer. Front Genet 14.

9. Cavic, M., Spasic, J., Krivokuca, A., Boljevic, I., Kuburovic, M., Radosavljevic, D., et al. (2019). TP53 and DNA-repair gene polymorphisms genotyping as a low-cost lung adenocarcinoma screening tool. J Clin Pathol 72, 75–80. doi: 10.1136/jclinpath-2018-205553.

10. Čavić. Milena, Krivokuća, A., Boljević, I., Brotto, K., Jovanović, K., Tanić. Miljana, et al. (2016). Pharmacogenetics in cancer therapy – 8 years of experience at the Institute for Oncology and Radiology of Serbia. J BUON 21, 1287–1295.

11. Cecchin, E., Agostini, M., Pucciarelli, S., De Paoli, A., Canzonieri, V., Sigon, R., et al. (2011). Tumor response is predicted by patient genetic profile in rectal cancer patients treated with neo-adjuvant chemo-radiotherapy. Pharmacogenomics J 11, 214–226. doi: 10.1038/tpj.2010.25.

12. Chandrashekar, D. S., Bashel, B., Balasubramanya, S. A. H., Creighton, C. J., Ponce-Rodriguez, I., Chakravarthi, B. V. S. K., et al. (2017). UALCAN: A Portal for Facilitating Tumor Subgroup Gene Expression and Survival Analyses. Neoplasia 19, 649–658. doi: 10.1016/j.neo.2017.05.002.

13. Choi, S., and Mason, J. (2000a). Choi SW, Mason JB. Folate and carcinogenesis: an integrated scheme. J Nutr 130, 129–132. J Nutr 130, 129–132.

14. Choi, S.-W., and Mason, J. B. (2000b). Folate and Carcinogenesis: An Integrated Scheme. J Nutr 130, 129–132. doi: 10.1093/jn/130.2.129.

15. De Mattia, E., Roncato, R., Palazzari, E., Toffoli, G., and Cecchin, E. (2020). Germline and Somatic Pharmacogenomics to Refine Rectal Cancer Patients Selection for Neo- Adjuvant Chemoradiotherapy. Front Pharmacol 11. doi: 10.3389/fphar.2020.00897.

16. Fekete, J. T., and Győrffy, B. (2019). ROCplot.org: Validating predictive biomarkers of chemotherapy/hormonal therapy/anti-HER2 therapy using transcriptomic data of 3,104 breast cancer patients. Int J Cancer 145, 3140–3151. doi: 10.1002/ijc.32369.

17. Garcia-Aguilar, J., Chen, Z., Smith, D. D., Li, W., Madoff, R. D., Cataldo, P., et al. (2011). Identification of a Biomarker Profile Associated With Resistance to Neoadjuvant Chemoradiation Therapy in Rectal Cancer. Ann Surg 254, 486–493. doi: 10.1097/SLA.0b013e31822b8cfa.

18. Gaunt, T. R., Rodriguez, S., Zapata, C., and Day, I. N. (2006). MIDAS: software for analysis and visualisation of interallelic disequilibrium between multiallelic markers. BMC Bioinformatics 7, 227. doi: 10.1186/1471-2105-7-227.

19. Glynne-Jones, R., Wyrwicz, L., Tiret, E., Brown, G., Rödel, C., Cervantes, A., et al. (2017). Rectal cancer: ESMO Clinical Practice Guidelines for diagnosis, treatment and follow-up. Annals of Oncology 28, iv22–iv40. doi: 10.1093/annonc/mdx224.

20. Gong, J.-M., Shen, Y., Shan, W.-W., and He, Y.-X. (2018). The association between MTHFR polymorphism and cervical cancer. Sci Rep 8, 7244. doi: 10.1038/s41598-018-25726-9.

21. Jakovljevic, K., Malisic, E., Cavic, M., Krivokuca, A., Dobricic, J., and Jankovic, R. (2012a). KRAS and BRAF mutations in Serbian patients with colorectal cancer. J BUON 17, 575–80.

22. Jakovljevic, K., Malisic, E., Cavic, M., Radulovic, S., and Jankovic, R. (2012b). Association between methylenetetrahydrofolate reductase polymorphism C677T and risk of chronic myeloid leukemia in Serbian population. Leuk Lymphoma 53, 1327– 1330.

23. Jiang, Q., Chen, K., Ma, X., Li, Q., Yu, W., Shu, G., et al. (2005). Diets, polymorphisms of methylenetetrahydrofolate reductase, and the susceptibility of colon cancer and rectal cancer. Cancer Detect Prev 29, 146–154. doi: 10.1016/j.cdp.2004.11.004.

24. Kawakami, K., Ruszkiewicz, A., Bennett, G., Moore, J., Watanabe, G., and Iacopetta, B. (2003). The folate pool in colorectal cancers is associated with DNA hypermethylation and with a polymorphism in methylenetetrahydrofolate reductase. Clin Cancer Res 9, 5860–5.

25. Kennedy, D. A., Stern, S. J., Matok, I., Moretti, M. E., Sarkar, M., Adams-Webber, T., et al. (2012). Folate Intake, MTHFR Polymorphisms, and the Risk of Colorectal Cancer: A Systematic Review and Meta-Analysis. J Cancer Epidemiol 2012, 1–24. doi: 10.1155/2012/952508.

26. Kim, J. C., Ha, Y. J., Roh, S. A., Cho, D. H., Choi, E. Y., Kim, T. W., et al. (2013). Novel Single-Nucleotide Polymorphism Markers Predictive of Pathologic Response to Preoperative Chemoradiation Therapy in Rectal Cancer Patients. International Journal of Radiation Oncology*Biology*Physics 86, 350–357. doi: 10.1016/j.ijrobp.2012.12.018.

27. Kim, Y., Pogribny, I., Basnakian, A., Miller, J., Selhub, J., James, S., et al. (1997). Folate deficiency in rats induces DNA strand breaks and hypomethylation within the p53 tumor suppressor gene. Am J Clin Nutr 65, 46–52. doi: 10.1093/ajcn/65.1.46.

28. Kim, Y.-I. (2003). Role of folate in colon cancer development and progression. J Nutr 133, 3731S–3739S. doi: 10.1093/jn/133.11.3731S.

29. Komlósi, V., Hitre, E., Pap, É., Adleff, V., Réti, A., Székely, É., et al. (2010). SHMT1 1420 and MTHFR 677 variants are associated with rectal but not colon cancer. BMC Cancer 10, 525. doi: 10.1186/1471-2407-10-525.

30. Kono, S., and Chen, K. (2005). Genetic polymorphisms of methylenetetrahydrofolate reductase and colorectal cancer and adenoma. Cancer Sci 96, 535–542. doi: 10.1111/j.1349-7006.2005.00090.x.

31. Krivokuca, A. M., Cavic, M. R., Malisic, E. J., Rakobradovic, J. D., Kolarevic-Ivankovic, D., Tomasevic, Z. I., et al. (2016). Polymorphisms in cancer susceptibility genes XRCC1, RAD51 and TP53 and the risk of breast cancer in Serbian women. Int J Biol Markers 31, e258-63. doi: 10.5301/jbm.5000201.

32. Landberg, T., Chavaudra, J., Dobbs, J., Gerard, J.-P., Hanks, G., Horiot, J.-C., et al. (2016a). Report 62. Journal of the International Commission on Radiation Units and Measurements os32, NP-NP. doi: 10.1093/jicru/os32.1.Report62.

33. Landberg, T., Chavaudra, J., Dobbs, J., Hanks, G., Johansson, K.-A., Möller, T., et al. (2016b). Report 50. Journal of the International Commission on Radiation Units and Measurements os26, NP-NP. doi: 10.1093/jicru/os26.1.Report50.

34. Leopardi, P. (2006). Effects of folic acid deficiency and MTHFR C677T polymorphism on spontaneous and radiation-induced micronuclei in human lymphocytes. Mutagenesis 21, 327–333. doi: 10.1093/mutage/gel031.

35. Levin, B. L., and Varga, E. (2016). MTHFR: Addressing Genetic Counseling Dilemmas Using Evidence-Based Literature. J Genet Couns 25, 901–911. doi: 10.1007/s10897-016-9956-7.

36. Lightfoot, T. J., Barrett, J. H., Bishop, T., Northwood, E. L., Smith, G., Wilkie, M. J. V., et al. (2008). Methylene Tetrahydrofolate Reductase Genotype Modifies the Chemopreventive Effect of Folate in Colorectal Adenoma, but not Colorectal Cancer. Cancer Epidemiology, Biomarkers & Prevention 17, 2421–2430. doi: 10.1158/1055-9965.EPI-08-0058.

37. Longley, D. B., Harkin, D. P., and Johnston, P. G. (2003). 5-Fluorouracil: mechanisms of action and clinical strategies. Nat Rev Cancer 3, 330–338. doi: 10.1038/nrc1074.

38. Matsuo, K., Ito, H., Wakai, K., Hirose, K., Saito, T., Suzuki, T., et al. (2005). One-carbon metabolism related gene polymorphisms interact with alcohol drinking to influence the risk of colorectal cancer in Japan. Carcinogenesis 26, 2164–2171. doi: 10.1093/carcin/bgi196.

39. Murtaugh, M. A., Curtin, K., Sweeney, C., Wolff, R. K., Holubkov, R., Caan, B. J., et al. (2007). Dietary intake of folate and co-factors in folate metabolism, MTHFR polymorphisms, and reduced rectal cancer. Cancer Causes & Control 18, 153–163. doi: 10.1007/s10552-006-0099-2.

40. National Cancer Institute and the National Human Genome Research Institute (n.d.). The Cancer Genome Atlas. Available at: https://www.cancer.gov/about-nci/organization/ccg/research/structural-genomics/tcga/studied-cancers [Accessed March 3, 2021].

41. Nielsen R, S. M. (2013). An Introduction to Population Genetics. Oxford University Press.

42. Nikas, J. B., Lee, J. T., Maring, E. D., Washechek-Aletto, J., Felmlee-Devine, D., Johnson, R. A., et al. (2015). A common variant in MTHFR influences response to chemoradiotherapy and recurrence of rectal cancer. Am J Cancer Res 5, 3231–40.

43. Nikolic, N., Radosavljevic, D., Gavrilovic, D., Nikolic, V., Stanic, N., Spasic, J., et al. (2021). Prognostic Factors for Post-Recurrence Survival in Stage II and III Colorectal Carcinoma Patients. Medicina (B Aires) 57, 1108. doi: 10.3390/medicina57101108.

44. Oken, M. M., Creech, R. H., Tormey, D. C., Horton, J., Davis, T. E., McFadden, E. T., et al. (1982). Toxicity and response criteria of the Eastern Cooperative Oncology Group. Am J Clin Oncol 5, 649–656. doi: 10.1097/00000421-198212000-00014.

45. Rai, V. (2015). Evaluation of the MTHFR C677T Polymorphism as a Risk Factor for Colorectal Cancer in Asian Populations. Asian Pac J Cancer Prev 16, 8093–8100. doi: 10.7314/apjcp.2015.16.18.8093.

46. Rai, V. (2016). Evaluation of the MTHFR C677T Polymorphism as a Risk Factor for Colorectal Cancer in Asian Populations. Asian Pacific Journal of Cancer Prevention 16, 8093–8100. doi: 10.7314/APJCP.2015.16.18.8093.

47. Ryan, B. M., and Weir, D. G. (2001). Relevance of folate metabolism in the pathogenesis of colorectal cancer. Journal of Laboratory and Clinical Medicine 138, 164–176. doi: 10.1067/mlc.2001.117161.

48. Ryan-Harshman, M., and Aldoori, W. (2007). Diet and colorectal cancer: Review of the evidence. Can Fam Physician 53, 1913–1920.

49. Salnikova, L. E., and Kolobkov, D. S. (2016). Germline and somatic genetic predictors of pathological response in neoadjuvant settings of rectal and esophageal cancers: systematic review and meta-analysis. Pharmacogenomics J 16, 249–265. doi: 10.1038/tpj.2015.46.

50. Serbian Cancer Registry (2022). Malignant tumours in republic of Serbia.

51. Sheng, X., Zhang, Y., Zhao, E., Lu, S., Zheng, X., Ge, H., et al. (2012). MTHFR C677T polymorphism contributes to colorectal cancer susceptibility: evidence from 61 case– control studies. Mol Biol Rep 39, 9669–9679. doi: 10.1007/s11033-012-1832-4.

52. Siddiqui, M. R. S., Bhoday, J., Battersby, N. J., Chand, M., West, N. P., Abulafi, A.-M., et al. (2016). Defining response to radiotherapy in rectal cancer using magnetic resonance imaging and histopathological scales. World J Gastroenterol 22, 8414– 8434. doi: 10.3748/wjg.v22.i37.8414.

53. Stanojevic, A., Samiotaki, M., Lygirou, V., Marinkovic, M., Nikolic, V., Stojanovic-Rundic, S., et al. (2023). Data independent acquisition mass spectrometry (DIA-MS) analysis of FFPE rectal cancer samples offers in depth proteomics characterization of response to neoadjuvant chemoradiotherapy. medRxiv.

54. Stojanovic-Rundic, S., Marinkovic, M., Cavic, M., Karapandzic, V. P., Gavrilovic, D., Jankovic, R., et al. (2021). The role of haematological parameters in predicting the response to radical chemoradiotherapy in patients with anal squamous cell cancer. Radiol Oncol 55, 449–458. doi: 10.2478/raon-2021-0039.

55. STRING database (n.d.). Available at: https://string-db.org [Accessed July 21, 2020].

56. Szklarczyk, D., Gable, A. L., Lyon, D., Junge, A., Wyder, S., Huerta-Cepas, J., et al. (2019). STRING v11: protein-protein association networks with increased coverage, supporting functional discovery in genome-wide experimental datasets. Nucleic Acids Res 47, D607–D613. doi: 10.1093/nar/gky1131.

57. Teng, Z., Wang, L., Cai, S., Yu, P., Wang, J., Gong, J., et al. (2013). The 677C.T (rs1801133) Polymorphism in the MTHFR Gene Contributes to Colorectal Cancer Risk: A Meta-Analysis Based on 71 Research Studies. PLoS One 8, e55332. doi: 10.1371/journal.pone.0055332.

58. Terrazzino, S., Agostini, M., Pucciarelli, S., Pasetto, L. M., Friso, M. L., Ambrosi, A., et al. (2006). A haplotype of the methylenetetrahydrofolate reductase gene predicts poor tumor response in rectal cancer patients receiving preoperative chemoradiation. Pharmacogenet Genomics 16, 817–824. doi: 10.1097/01.fpc.0000230412.89973.c0.

59. The Human Protein Atlas database V.20.0 (n.d.). Available at: http://www.proteinatlas.org/pathology [Accessed March 3, 2021].

60. Thomas, F., Motsinger-Reif, A. A., Hoskins, J. M., Dvorak, A., Roy, S., Alyasiri, A., et al. (2011). Methylenetetrahydrofolate reductase genetic polymorphisms and toxicity to 5- FU-based chemoradiation in rectal cancer. Br J Cancer 105, 1654–1662. doi: 10.1038/bjc.2011.442.

61. Tong, W., Tong, G., Jin, D., and Lv, Q. (2018). MTHFR C677T and A1298C polymorphisms and lung cancer risk in a female Chinese population. Cancer Manag Res 10, 4155–4161. doi: 10.2147/CMAR.S176263.

62. UALCAN database (n.d.). Available at: http://ualcan.path.uab.edu/analysis.html [Accessed July 21, 2020].

63. Uhlen, M., Zhang, C., Lee, S., Sjöstedt, E., Fagerberg, L., Bidkhori, G., et al. (2017). A pathology atlas of the human cancer transcriptome. Science 357. doi: 10.1126/science.aan2507.

64. (UICC), U. for I. C. C. (2020). The TNM Classification of Malignant Tumours 8th edition. Available at: https://www.uicc.org/sites/main/files/atoms/files/UICC TNM Classification 8 ed. Errata 6 October 2020.pdf [Accessed March 11, 2021].

65. Ulrich, C. M., Rankin, C., Toriola, A. T., Makar, K. W., Altug-Teber, Ö., Benedetti, J. K., et al. (2014). Polymorphisms in folate-metabolizing enzymes and response to 5- fluorouracil among patients with stage II or III rectal cancer (INT-0144; SWOG 9304). Cancer 120, 3329–3337. doi: 10.1002/cncr.28830.

66. Vecchio, F. M., Valentini, V., Minsky, B. D., Padula, G. D. A., Venkatraman, E. S., Balducci, M., et al. (2005). The relationship of pathologic tumor regression grade (TRG) and outcomes after preoperative therapy in rectal cancer. International Journal of Radiation Oncology*Biology*Physics 62, 752–760. doi: 10.1016/j.ijrobp.2004.11.017.

67. Vuletić, A., Mirjačić Martinović, K., Tišma Miletić, N., Zoidakis, J., Castellvi-Bel, S., and Čavić, M. (2021). Cross-Talk Between Tumor Cells Undergoing Epithelial to Mesenchymal Transition and Natural Killer Cells in Tumor Microenvironment in Colorectal Cancer. Front Cell Dev Biol 9. doi: 10.3389/fcell.2021.750022.

68. Wang, J., Gajalakshmi, V., Jiang, J., Kuriki, K., Suzuki, S., Nagaya, T., et al. (2006a). Associations between 5,10-methylenetetrahydrofolate reductase codon 677 and 1298 genetic polymorphisms and environmental factors with reference to susceptibility to colorectal cancer: A case-control study in an Indian population. Int J Cancer 118, 991–997. doi: 10.1002/ijc.21438.

69. Wang, J., Gajalakshmi, V., Jiang, J., Kuriki, K., Suzuki, S., Nagaya, T., et al. (2006b). Associations between 5,10-methylenetetrahydrofolate reductase codon 677 and 1298 genetic polymorphisms and environmental factors with reference to susceptibility to colorectal cancer: A case-control study in an Indian population. Int J Cancer 118, 991–997. doi: 10.1002/ijc.21438.

70. Wei, J., Huang, R., Guo, S., Zhang, X., Xi, S., Wang, Q., et al. (2018). ypTNM category combined with AJCC tumor regression grade for screening patients with the worst prognosis after neoadjuvant chemoradiation therapy for locally advanced rectal cancer. Cancer Manag Res 10, 5219–5225. doi: 10.2147/CMAR.S179151.

71. Yamagishi, H., Kuroda, H., Imai, Y., and Hiraishi, H. (2016). Molecular pathogenesis of sporadic colorectal cancers. Chin J Cancer 35, 4. doi: 10.1186/s40880-015-0066-y.

72. Yin, G., Kono, S., Toyomura, K., Hagiwara, T., Nagano, J., Mizoue, T., et al. (2004). Methylenetetrahydrofolate reductase C677T and A1298C polymorphisms and colorectal cancer: The Fukuoka Colorectal Cancer Study. Cancer Sci 95, 908–913. doi: 10.1111/j.1349-7006.2004.tb02201.x.

73. Zhao, M., Li, X., Xing, C., and Zhou, B. (2013). Association of methylenetetrahydrofolate reductase C677T and A1298C polymorphisms with colorectal cancer risk: A meta-analysis. Biomed Rep 1, 781–791. doi: 10.3892/br.2013.134.

74. Zhao, Y., Li, X., and Kong, X. (2015). MTHFR C677T Polymorphism is Associated with Tumor Response to Preoperative Chemoradiotherapy: A Result Based on Previous Reports. Medical Science Monitor 21, 3068–3076. doi: 10.12659/MSM.895433.

